# Assortative social mixing and sex disparities in tuberculosis burden

**DOI:** 10.1101/2020.11.18.20233809

**Authors:** Debebe Shaweno, Katherine Horton, Richard Hayes, Peter J. Dodd

## Abstract

Globally, men have higher tuberculosis (TB) burden but the mechanisms underlying this sex disparity are not fully understood. Recent surveys of social mixing patterns have established moderate preferential within-sex mixing in many settings. This assortative mixing could amplify differences from other causes. We explored the impact of assortative mixing and factors differentially affecting disease progression and detection using a sex-stratified deterministic TB transmission model. We explored the influence of assortativity at disease-free and endemic equilibria, finding stronger effects during invasion and on increasing male:female prevalence (M:F) ratios than overall prevalence. Variance-based sensitivity analysis of endemic equilibria identified differential progression as the most important driver of M:F ratio uncertainty. We fitted our model to prevalence and notification data in examplar settings within a fully Bayesian framework. For our high M:F setting, random mixing reduced equilibrium M:F ratios by 12%(95%CrI 0 - 30%). Equalizing male case detection there led to a 20% (95%CrI 11 - 31%) reduction in M:F ratio over 10 years - insufficient to eliminate sex disparities. However, this potentially achievable improvement was associated with a meaningful 8%(95%CrI 4 - 14%) reduction in total TB prevalence over this time frame.

## Introduction

Considerable sex disparity exists in the burden of tuberculosis (TB) across all regions of the world. Global estimates indicate that around 64% of all TB incidence in adults (aged ≥15 years) is among men.^1^ The ratio of men-to-women for prevalent bacteriologically confirmed pulmonary TB shows wide geographic variations with ratios ranging from 1.2 in Ethiopia to about 5 in Vietnam.^1^ Similarly, national TB prevalence surveys have found male-to-female ratios in prevalent bacteriologically-confirmed TB (M: F ratios) of around 2.2 across low- and middle-income countries.^2^ Even in settings with generalised HIV epidemics, where the prevalence of HIV (a strong risk factor for TB) is higher among women than men, men have higher TB prevalence than women.^2^

The underlying reasons behind excess male burden in different measures are not fully understood. Some studies based on routinely diagnosed TB cases have suggested that the excess notifications among men is a result of systematic underdiagnosis or under reporting of TB in women due to differences in access to care between women and men.^3^ However, prevalence surveys from different settings have usually shown not only a male dominance among undiagnosed cases, but also higher prevalence-to-notification ratios for men, implying a male disadvantage in accessing TB care.^2,4^ In addition, observations that sex disparities in TB rates do not arise before the age of puberty^3,5^ have led to speculations around the role of immunological and behavioural factors.^6,7^ Several studies have established a link between TB and behaviours such as smoking and alcohol consumption^8,9^ which are more frequent in men than in women in many high TB burden settings,^1,6,10^ and which are known to enhance the risk of TB progression and transmission.^11,12^

Sex differences in social roles between men and women can determine the patterns and location of social mixing relevant to transmission of *Mycobacterium tuberculosis* (*M.tb*).^7^ Previous studies have linked substantial TB incidence in adults to their preferential contact with other adults.^13^ It has been estimated that 80-90 percent of TB infections are acquired from social contacts occurring outside households.^14–16^ Many of these infections likely occur in congregate settings which enhance sex-assortative social mixing such as homeless shelters, prisons, public transports, and bars.^17,18^ Consistent with these reports, a recent systematic review of social contact patterns found widespread sex-assortative social mixing, and also higher rates of workplace contact for men.^19^ Modelling of the implications of social contact data from Zambia and South Africa for *M.tb* forces-of-infection suggested higher forces-of-infection among men due to sex-assortative mixing and higher male TB prevalence.^13^ Sex-assortative mixing therefore provides a mechanism that could amplify disparities in TB burden between men and women arising from other underlying factors.

In this study, we use a sex-stratified TB transmission model to assess the relative role of social mixing, differential progression, and differential case detection rates in establishing and strengthening sex disparities in TB prevalence. Bayesian calibration of our model to two exemplar settings allows us to characterize parameter regions compatible with observations, and assess whether the residual uncertainties affect predicted intervention impact.

## Methods

### Conceptual framework, model structure, and priors

We use a conceptual framework similar to that developed for considering other social determinants of TB,^20^ and implement these mechanisms within a standard compartmental transmission model structure for adults over 15 years of age (see Figure 1). We consider three types of mechanism by which men may come to have a higher TB prevalence than women: 1) risk factors directly affecting the natural history of infection, progression and relapse; 2) risk factors that lead to worse care access and therefore longer duration of TB disease among men; 3) risk factors relating to transmission patterns. Data are lacking to support sex-specific mortality and self-cure from untreated TB,^21^ and so we did not include this possibility.

**Figure 1.**
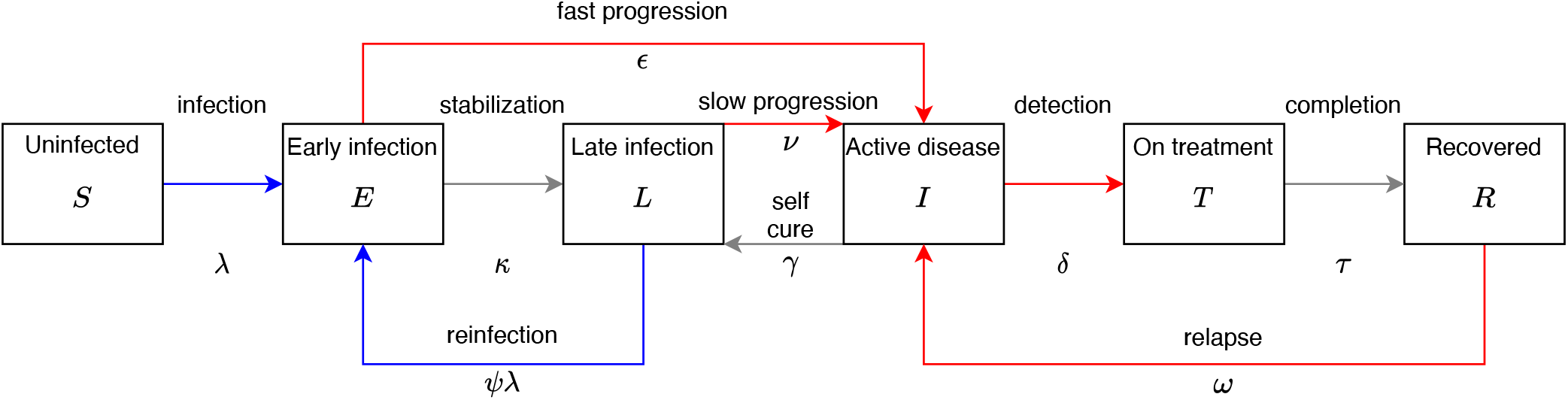
Model structure, duplicated for each sex. Red transitions are sex-dependent to represent different risks in disease progression and care access. Blue transitions represent infection, and are also sex-dependent due to assortative mixing. Not shown: death from all states at rate *µ* (and at an additional rate *µ*_*t*_ from active TB disease); birth into Uninfected, at rate to keep population fixed.

A key interest for us is the role of assortative mixing between men and women. We model this using a single parameter that interpolates between random mixing between the sexes, and no mixing between different sexes, without changing the total contact rates (assumed the same for men and women). Natural history risk aspects affected may include increased risk of infection, increased risk of progression following (re)infection, as well as increased risk of relapse. Causes may include immunological differences between sexes as well as different levels of modifiable proximal risk TB factors such as smoking. We model these as a hazard ratio for progression or relapse, ie. we modelled differential fast progression for men as *ε*_*m*_ = *α* × *ε/*(1 + *α*) and for women as *ε* _*f*_ = *ε/*(1 + *α*), so that the average progression parameter, *ε*, can be informed by literature that does not stratify by sex. We modelled the sex-specific rates of reactivation (*ν*) and relapse (*ω*) analogously, using the same parameter *α*. The prior for factor *α* is chosen based on genotypic studies that compared reactivation rates between men and women,^22^ but increased the reported standard deviation to 1 to allow for greater differences in other settings: see Table 1. Worse access to care may reflect differences in recognition and response to illness, barriers in accessing care, and health systems features; these are modelled in the same was as differential progression, as a hazard ratio in moving from symptomatic disease to treatment.

**Table 1.**
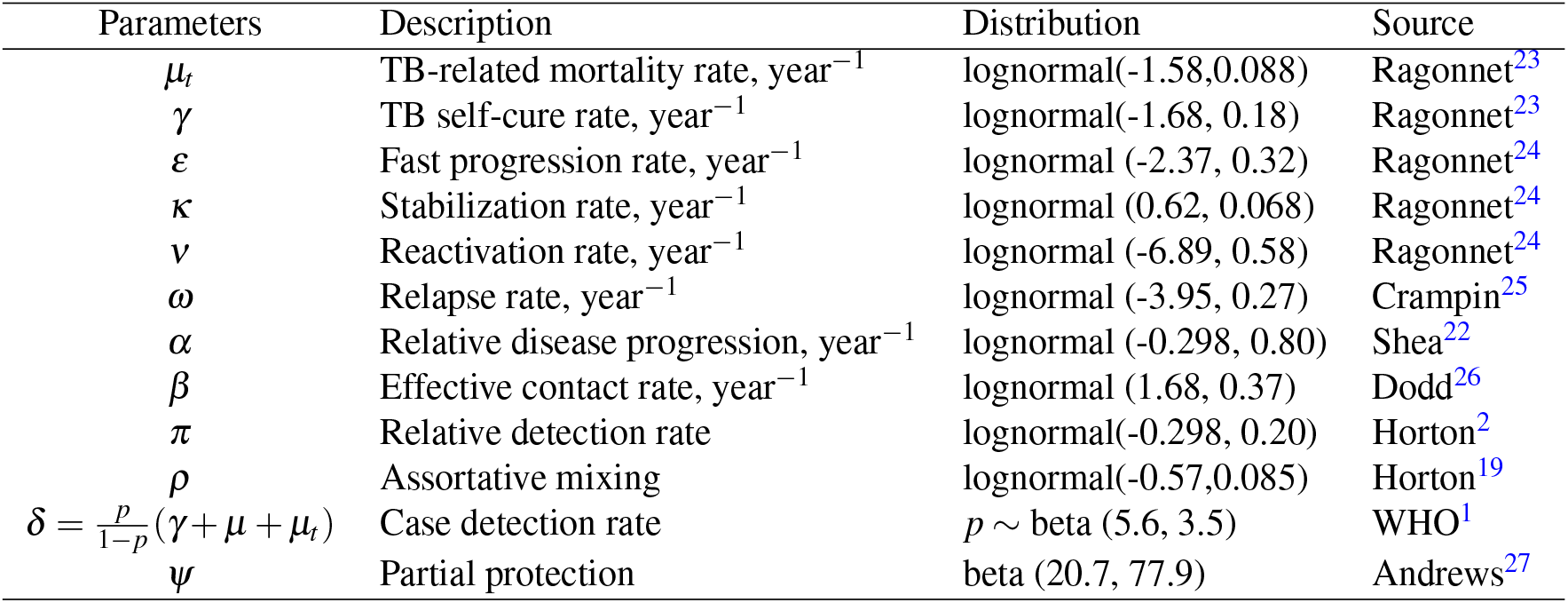
Priors and sources for model parameters. NB the prior for *δ* is defined by comparison to competing hazard parameters to give an uninformative prior on the probability of a TB case being notified (*p*). We chose the *α* prior so that the mid value is centered at 1.0^22^ and increased the reported standard deviation to 1 to allow for greater differences in other settings.

We modelled a fixed population size with births equal to deaths, and half of the population being male at all times. With _*i*_ = *m, f* for men and women, respectively, the ordinary differential equations for this model are given in Equations 1 and 2.

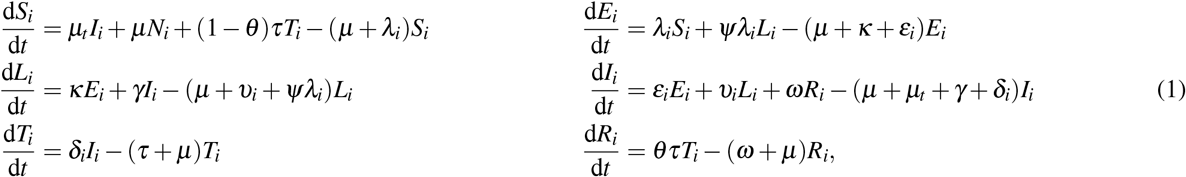

Here, the states and dates are defined in Figure 1. The total populations are *N*_*i*_ = *S*_*i*_ + *E*_*i*_ + *L*_*i*_ + *I*_*i*_ + *T*_*i*_ + *R*_*i*_. A fraction (1 *−θ*) of those on anti-TB treatment are assumed to die. The forces-of-infection in men and women are given by

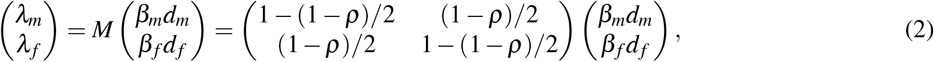

where *d*_*i*_ = *I*_*i*_*/N*_*i*_. We also consider differential infectiousness by sex as *β*_*m*_ = *f*_*m*_× *β* and *β*_*f*_ = *f* _*f*_ × *β* where *f*_*m*_ and *f* _*f*_ are the fractions of prevalent TB cases that are sputum smear positive among men and women, respectively. The assortativity parameter *ρ* in the mixing matrix *M* ranges from −1 (mixing only with the opposite sex), through 0 (representing random mixing), to 1 (completely within-sex mixing). The proportion, *m*, of effective contacts each sex makes with the same sex is given by *m* = (*ρ* + 1)*/*2. Below, we will refer to the one-sex model, which is defined by considering one of *i* = *m, f* separately with *ρ* = 1 (so that *λ*_*i*_ = *β*_*i*_*d*_*i*_). Table 1 defines the priors we use for our model parameters, and the sources we base them on.

### Reproduction number and invasion dynamics

In order to explore the impact of mixing on the early dynamics of TB within a susceptible population (invasion) for our model, we analytically calculate *R*_0_ and the M:F ratio during the associated exponential growth phase from the disease-free equilibrium. We calculate how these metrics vary with assortativity at different levels of other parameters.

### Equilibria and sensitivity analysis

Similarly, we numerically compute the equilibrium TB prevalence and M:F ratio by running the model for 400 years, and explore how these vary with assortativity. To robustly characterise the importance of different parameters given our priors, including their interactions, we calculated the Sobol’ variance-based sensitivity indices^28^ for the outcomes of total TB prevalence and M:F ratio. We used the SALib Python library,^29^ generating 2000 parameter sets using the priors detailed in Table 1. Code for all analyses is available at https://github.com/Debebe/tbsex.

### Approach to calibration

We used Mathematica 12.1^30^ to obtain quadratic equation for the one-sex model equilibrium prevalence by setting the left hand side of Equations 1 to zero. This was of the form

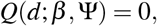

where *d* = *D/N*, and Ψ represents the parameters other than *β*. The two-sex model has identical form for men and women (with different parameter values); the dynamics are coupled only through the force of infection in Equation 2, with *λ*_*i*_, *i* = *m, f*, playing the role of *βd* in the one-sex model. This means the condition for the two-sex model to be at equilibrium can be written

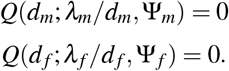

Approximate inference can then be performed using the log-likelihood

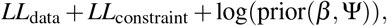

where

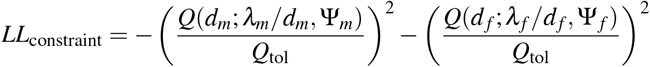

ensures that the prevalences are close to that generated at equilibrium under the input parameters (to within a tolerance *Q*_*tol*_). Smaller values of *Q*_*tol*_ result in sampled prevalences that are more tightly peaked around equilibrium values implied by parameters. We used a value of *Q*_*tol*_ = 10^*−*6^.

The log-prior is the sum over the individual log-priors in Table 1. The data likelihood is given by

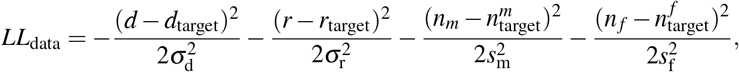

where *d* and *r* are the overall prevalence and M:F ratios, respectively, and *n*_*i*_ are the sex-specific notification rates. For Uganda, the observed values of *d* and *r* are 401 per 100,000 and 4.1 respectively^31^, while for Ethiopia the corresponding values were 277 per 100,000 and 1.2.^32^ We used the *n*_*m*_ and *n* _*f*_ values of respectively 161 and 79 per 100,000 for Uganda^31^, and 208 and 169 per 100,000 for Ethiopia^1^ (see Table 2). We assumed the standard deviations were 10% of their corresponding targets, i.e. 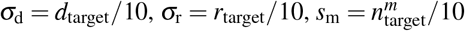 and 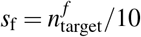.

**Table 2.** Posterior estimates for exemplar settings: calibration targets compared to data, and parameters determining sex difference for TB.

|  | Ethiopia |  | Uganda |  |
| --- | --- | --- | --- | --- |
|  | Data | Posterior (95% CrI) | Data | Posterior (95% CrI) |
| Prevalence | 277 (208, 347)<sup>32</sup> | 283 (232, 336) | 401(292, 509)<sup>31</sup> | 397 (321, 474) |
| Prevalence M:F ratio | 1.2<sup>32</sup> | 1.24 (1.03, 1.45) | 4.1<sup>31</sup> | 3.76 (3.0, 4.5) |
| Notifications | 188 | 181 (155,208) | 119<sup>31</sup> | 120 (103, 136) |
| Notifications M:F ratio | 1.2 | 1.11(0.87, 1.40) | 2.03<sup>31</sup> | 2.23 (1.75, 2.86) |
| Detection M:F ratio ($\pi$) | - | 0.90 (0.69, 1.16) | - | 0.60 (0.46, 0.78) |
| Progression M:F ratio ($\alpha$) | - | 1.12 (0.95, 1.32) | - | 2.75 (2.17, 3.45) |
| Assortativity ($\rho$) | - | 0.14 (-0.04, 0.34) | - | 0.16 (-0.02, 0.36) |

Calibration was performed using no-U-turn Hamiltonian Markov chain Monte Carlo using Stan.^33^ Three chains were run for 10,000 iterations and the first 5,000 runs were excluded as burn-in.

### Exemplar settings and data

We chose one setting with high M:F ratio (Uganda) and one with low M:F ratio (Ethiopia). For these exemplar settings, we extracted data on sex-stratified bacteriologically-confirmed TB prevalence from national prevalence surveys of Uganda (2014-2015)^34^ and Ethiopia (2010-2011).^32^ In both settings, the proportions prevalent with smear-positive disease (which we used to parametrize relative infectiousness) were higher in men: *f*_*m*_ = 0.45 vs *f* _*f*_ = 0.35 in Ethiopia,^32^ and *f*_*m*_ = 0.43 vs *f*_*m*_ = 0.38 in Uganda.^31^ We also compiled notification and case detection rates from WHO reports on the same year of prevalence surveys in those settings.^1^

Sex-specific prevalence and notification rates for each setting were used as calibration targets (see previous section). All data used in this study represent only individuals older than 15 years. For Ethiopia, sex-stratified notifications for bacteriologically confirmed TB were not available in the year of prevalence survey (2011). We therefore generated male-to-female ratios for new and relapse TB notifications from the closest year (i.e., 2013), and applied these values to generate sex-specific bacteriologically confirmed notification calibration targets for 2011. Models for the two exemplar settings were identical and used the same prior distributions (Table 1), but used setting-specific values for background mortality (from United Nations World Population Prospects estimates) and anti-TB treatment success (from World Health Organization collated treatment outcome data).

### Relevance to interventions; contribution of assortativity to prevalence M:F ratios

We simulated an intervention (eliminating the sex gap in access to care) and evaluated the relative impact on TB M:F ratio and prevalence after 10 years. We reported posterior mean and 95% quantiles of percentage reductions for each setting. Anticipating non-identifiability and potential posterior correlations between assortativity and other parameters influencing the M:F ratio, we assessed whether different combinations of parameters with similar posterior probability resulted in different dynamics under interventions. We selected three pairs of parameters that generate sex differences for TB: *α*, determining the relative risk of progression; *π*, determining the relative detection rate; and *ρ*, determining the assortativity of social mixing. For each parameter pair we drew an elliptical region with major and minor radii of respectively 0.05 and 0.02 on their joint posterior distributions. The ellipses were centered at the 25, 50 and and 75th quantiles for each variable, and were labelled respectively ‘L’ for low, ‘M’ for middle, and ‘U’ for upper. We used all samples with in ellipses for parameter pairs of interest and random samples of all other parameters to simulate the the impact of intervention. The number of parameters within each ellipse varied between 90-422 depending on sampled region, parameter combination and the exemplar setting.

To assess the contribution of assortativity to M:F ratios in these settings, we evaluated the mean percentage reduction in equilibrium M:F ratio (and 95% quantiles) from moving to a counterfactual with random mixing, across 200 samples from the full posterior.

## Results

### Influence of assortativity on invasion dynamics and equilibria

#### The one-sex model

For a compartmental TB transmission model such as this, we found the mean number of secondary cases generated at disease free equilibrium (*R*_0_) as *R*_0_ = (infectiousness) × (mean duration) × (progression probability) = *β* × *T*× *P*. For TB models the structure of Equation 1, there are two routes of progression from infection to active disease - fast and slow - so that:

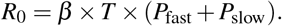

In our model,

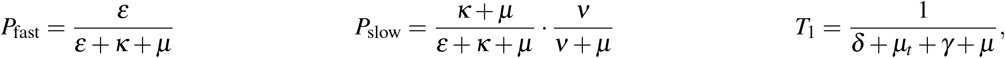

where *T*_1_ is the mean duration of a single TB episode. In reality (and our model) multiple episodes of TB are possible due to relapse and self-cure followed by reactivation. The probability of each subsequent episode was therefore the sum over these routes, *P*_+_ = *P*_self-cure_ + *P*_relapse_, and the mean time with TB, *T*, given as a sum over each possible number of episodes:

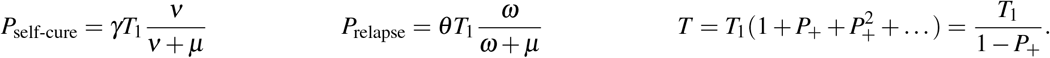

These formulae were used to compute *R*_*m*_ and *R*_*f*_ - the values of *R*_0_ for a one-sex model with parameters for men and women, respectively.

#### The two-sex model

For the two-sex model, *R*_0_ was calculated as the largest eigenvalue of the next generation matrix *G*, given by

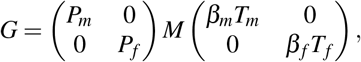

where *M* is the mixing matrix defined in Equation 2, and *P*_*m*_, *β*_*m*_, and *T*_*m*_ are the values of *P, β*, and *T* for men (and similarly for women, with subscript *f*). This was solved exactly to give

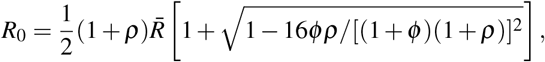

where 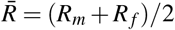 and *φ* = *R*_*m*_*/R*_*f*_. Solving for the corresponding eigenvector allowed calculation of the M:F incidence ratio during exponential growth as a function of *ρ*, which was found to satisfy

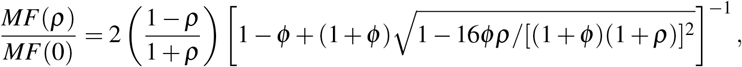

where *MF*(0) = *P*_*m*_*/P*_*f*_. Thus, for a given 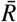 and *MF*(0) (respectively), *R*_0_ and the early M:F incidence ratio only depend on assortativity (*ρ*) and the ratio of male:female one-sex *R*_0_ values (*φ*). The behaviour of *R*_0_ and *MF*(*ρ*) as assortativity varies is shown for various choices of *φ* in Figure 2, panels a) and b).

**Figure 2.**
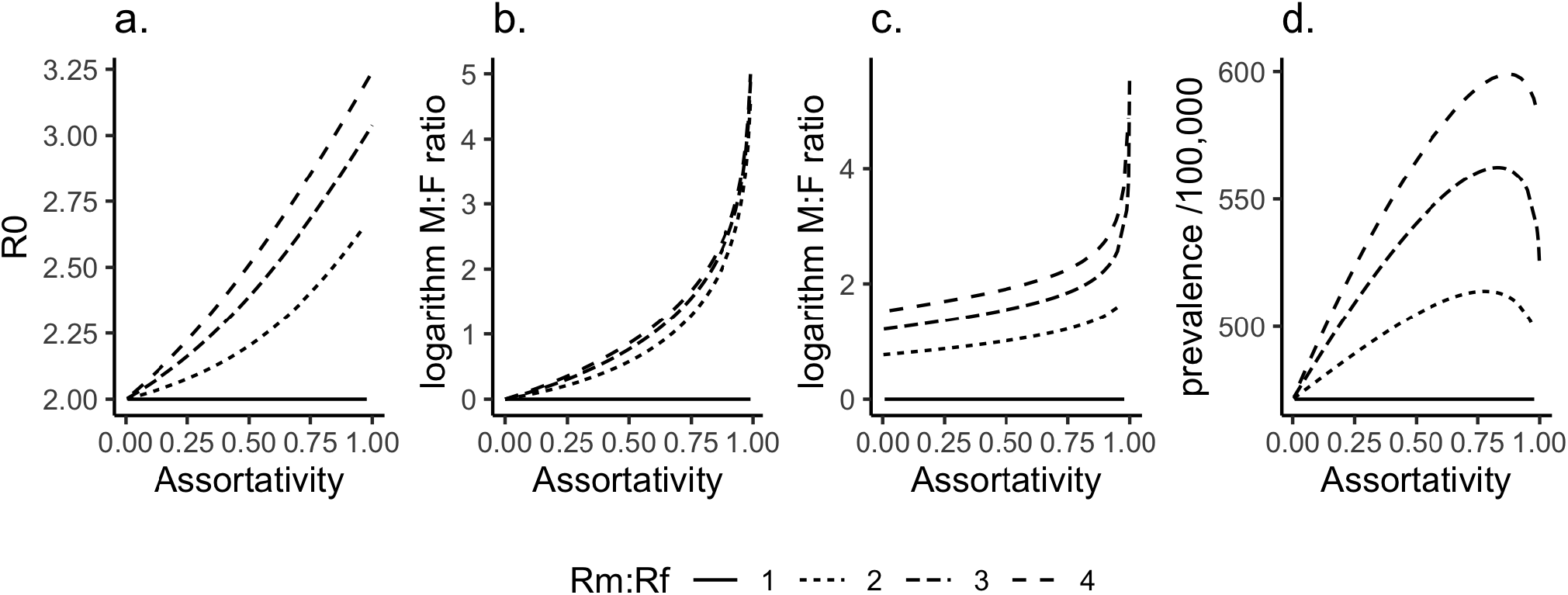
Influence of assortativity (*ρ*) at invasion and equilibria for different *R*_*m*_*/R*_*f*_. The effect of *ρ* on: a) *R*_0_; b) M:F ratio during early exponential growth from the disease-free equilibrium; c) M:F ratio in TB prevalence at endemic equilibrium; d) TB prevalence at endemic equilibrium. In c) and d), *β* and *α* were solved to give *R*_0_ = 2 and fix *R*_*m*_*/R*_*f*_, *ρ* was varied, and *π* = 1. Other parameters were the means of priors in Table 1.

#### Equilibria

The corresponding changes in equilibrium TB prevalence and equilibrium M:F ratio are shown in Figure 2, panels c) and d). Analytical solution was not possible, and (unlike the invasion dynamics) these results may depend on the particular parameter combinations that were used. Parameter values corresponding to the means of priors in Table 1 were used, except for: *ρ*, which was varied between 0 and 1; *β* and *α*, which were solved to fix *R*_0_ = 2 and various *R*_*m*_*/R*_*f*_; and *π*, which was set to 1 to allow *R*_*m*_*/R*_*f*_ to attain a value of 1.

### Sensitivity of all parameters’ influence on equilibria

At equilibrium the variance in TB M:F ratio is most influenced by a single model parameter – differential progression *α*, see Figure 3. The first- order effect of *α* on TB M:F ratio is 0.79. On the other hand, the differential detection parameter, *π* and the mixing parameter (*m* = (*ρ* + 1)*/*2 being the proportion of effective contacts that are same sex) have first order effects of 0.01 and 0.009 respectively. All other model parameters have first-order effects less than 0.009. By interacting with other model parameters: *α, m, p, β* and *π* attained total-order effects of 0.96, 0.04, 0.03, 0.03 and 0.02 respectively.

**Figure 3.**
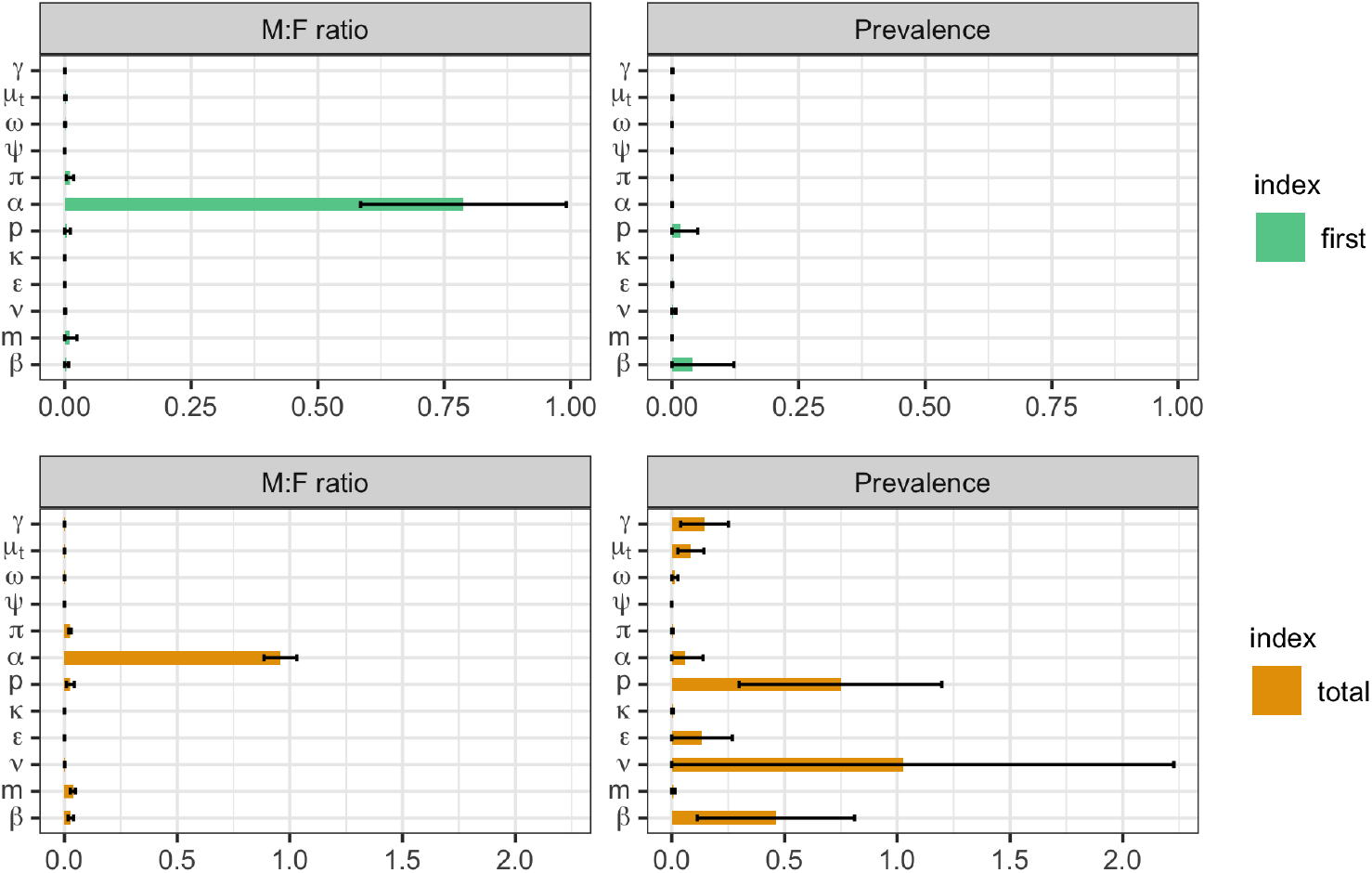
Sobol’ first-order (one-way) and total sensitivity indices (including interactions) for equilibrium prevalence M:F ratio and total TB prevalence (shown on x-axes). Distributions of parameters (y-axis) are as in Table 1 (*Ψ* - partial protection; *ν* - reactivation; *ε* - fast progression; *κ* - stabilization; *ω* - relapse; *γ* - self-cure; *µ*_*t*_ - TB mortality; *p* - detection; *β* - effective contact; *ρ* = 2*m−* 1 - assortativity; *π* - differential detection; *α* - differential progression).

For equilibrium TB prevalence, the main influence comes from reactivation rate (*ν*), detection rate (*δ*), effective contact rate (*β*), and fast progression rate (*ε*). Their total effects were attained by their interaction with other parameters (their sum is *≥* 1).

### Calibration to exemplar settings

Explicit formulae for the quadratic function *Q*(*d, β*; Ψ) used in fitting are given in the Appendix, as are example Markov chain plots of samples. The Gelman-Rubin convergence diagnostic statistic exhibited 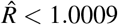 for all model parameters. For each exemplar setting, the fits to calibration target data and estimates of the parameters modelling sex disparities are shown in Table 2. The smoothed pairs plot for the 8-parameter posterior in Ethiopia is shown in Figure 4.

**Figure 4.**
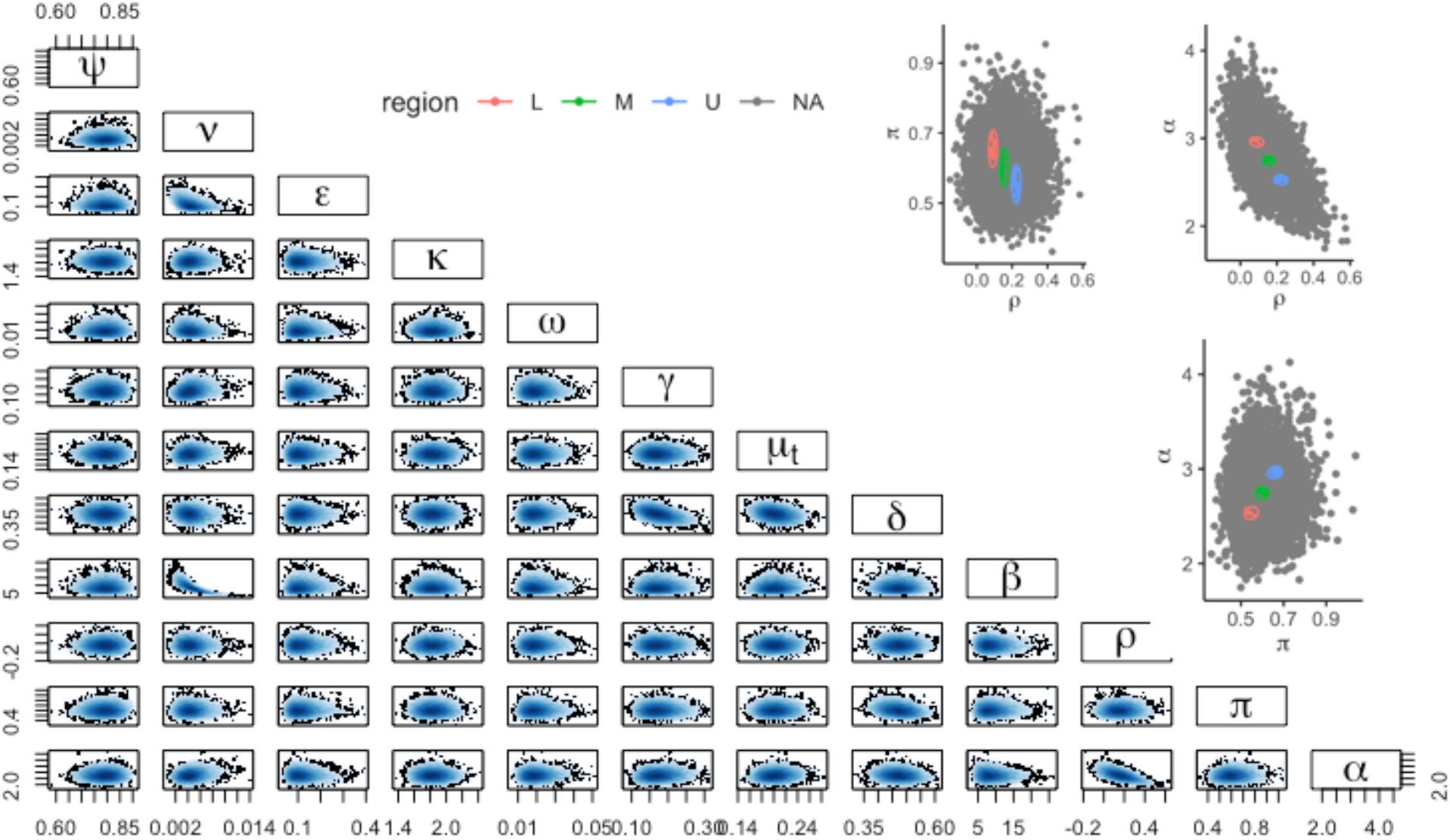
Correlation between model parameters. The lower left corner plot shows the smoothed posterior samples (*Ψ* - partial protection; *ν* - reactivation; *ε* - fast progression; *κ* - stabilization; *ω* - relapse; *γ* - self-cure; *µ*_*t*_ - TB mortality; *p* - detection; *β* - effective contact; *ρ* - assortativity; *π* - differential detection; *α* - differential progression). The upper right plots show the ‘L’, ‘M’, and ‘U’ ellipses used for the sensitivity analysis in Table 3.

### Intervention results; contribution of assortativity

Sampling across the whole posterior for each setting, we found the hypothetical intervention reduced TB prevalence after 10 years in comparison to the counterfactual of no intervention by 0.7% (95%CrI −0.3 - 2.7%) for Ethiopia and 8% (95%CrI 4 - 14%) for Uganda. The M:F ratios reduced over the same time time period by 7% (95%CrI −10 - 21%) for Ethiopia and 20% (95%CrI 11 - 31%) for Uganda. The 3 elliptical regions selected for each parameter pair (*π, α*), (*ρ, α*), (*ρ, π*) in order to assess the impact of remaining uncertainty on projected intervention impacts are shown in Figure 4 (top right three panels). Comparisons of posterior log-likelihoods for these regions are shown in the Appendix, alongside time series plots of prevalence under the intervention. The percentage reductions in TB prevalence and M:F ratio for each setting, parameter pair, and L, M, and U sampling regions are given in Table 3.

**Table 3.** Sensitivity analysis of percentage reduction in TB prevalence and M:F ratios sampling from restricted regions of the posterior. ‘L’, ‘M’, and ‘U’ correspond to different elliptical regions shown top-right in Figure 4. (*ρ* - assortativity; *π* - differential detection; *α* - differential progression).

| Outcome | Parameter pair | Ethiopia |  |  | Uganda |  |  |
| --- | --- | --- | --- | --- | --- | --- | --- |
|  |  | L | M | U | L | M | U |
| Prevalence | $\pi, \alpha$ | 0.9(0.5,1.4) | 0.5(0.3,0.8) | 0.1(-0.04, 0.2) | 10(7,15) | 9(6,11) | 7(5,9) |
| | $\rho, \alpha$ | 0.8(-0.3,3) | 0.6(-0.3,3) | 0.6(-0.2,0.3) | 9(4,15) | 8(3, 13) | 8(3,13) |
| | $\rho, \pi$ | 0.9(0.3,1.8) | 0.4(0.04,1.1) | 0.1(0,0.3) | 10(7,13) | 9(6, 12) | 7(5, 9) |
| TB M:F ratio | $\pi, \alpha$ | 11(9,14) | 6(5,8) | 1(-0.5,3) | 24(20,27) | 21(17,24) | 17(14,20) |
| | $\rho, \alpha$ | 6(-9,21) | 5(-11,19) | 6(-9,21) | 21(12,30) | 20(8, 29) | 20(9,29) |
| | $\rho, \pi$ | 11(8,14) | 6(4,9) | 1.5(-1.3, 4) | 23(19,28) | 20(17,24) | 18(14,21) |

Sampling across the whole posterior for each setting, we found equilibria with random mixing had a prevalence M:F ratio that was 12%(−0.2 - 30%) lower in Uganda. For Ethiopia, equilibria with random mixing has equilibrium prevalence M:F ratio lower by 3%(−0.5 - 10%) compared to equilibrium M:F ratio based on assortativity parameter sampled from the whole region.

## Discussion

Using a TB transmission dynamic model and Bayesian inference framework, we determined the extent to which social mixing and other factors contribute to sex disparity in TB prevalence. Our analysis shows that sex disparity in TB burden results from a complex interplay among several factors that determine progression, detection and social mixing, although different factors dominate at different stages of the epidemic. The impact of assortativity is much larger at invasion compared to its role at endemic equilibrium. Our fit to a high M:F prevalence ratio setting (Uganda) suggested 12%(0 - 30%) of the M:F ratio could be attributed to the effects of assortative mixing.

Tuberculosis invades more rapidly as assortative sex mixing increases because men are more likely to transmit infection to other men due to the more frequent interactions with males^19^ as well as due to higher burden of infectious TB among men.^32,35^ Men have higher rates of mixing with other men meaning that they can generate more secondary cases preferentially among men. As a result, early growth sex disparity in TB burden between men and women is extremely wide. The importance of assortativity during invasion dynamics has relevance to TB outbreaks, and potentially to invasion and replacement by new strains (eg. drug-resistant strains) in TB-endemic settings.

At invasion, higher assortative mixing increases overall TB prevalence. At endemic equilibrium, increasing assortative mixing leads to a gradual initial rise in an overall equilibrium prevalence followed by a subsequent fall as assortativity approaches 1 (Figure 2c). The peak represents a balance between the amplifying effects of increasing assortativity, and the folding in of infections among only the half of the population at higher risk (men) as assortativity approaches 1.

Although, we found lower male case detection rates in Uganda (our high M:F ratio setting), our finding for Ethiopia (our low M:F ratio setting) is different. In Ethiopia, case detection rates between men and women are nearly comparable, with empirical prevalence-to-notification ratios of 1.7 and 1.5, respectively – implying no considerable differences between men and women in accessing TB care. These findings are consistent with some studies,^36^ and explain the limited impacts of our hypothetical intervention in this setting.

Interventions targeting only differential case detection are unlikely to eliminate sex disparities in TB burden alone. We found closing disparities in detection in a high M:F ratio setting (Uganda) would reduce the M:F ratio by around 20% over 10 years, but would also reduce TB prevalence by around 8%. Elimination of sex disparities in TB burden will therefore also require interventions targeting modifiable risk factors that are more common among men.

We found the predicted impact of narrowing case detection gaps was not strongly dependent on posterior parameter region in our high M:F ratio setting. This is reassuring, in that while data does not completely identify the exact contribution of different mechanisms to observed sex disparities in TB, our ignorance need not serve as a barrier to projecting the likely impact of sex-specific interventions. For Ethiopia, with only a small sex difference in baseline detection rates, the exact size of this differential did influence projected impact on M:F ratio, but not appreciably the impact TB prevalence.

A strength of this study was our inference approach, which treated all model parameters as uncertain, specified via priors, within a Bayesian framework. This was necessary in order to answer our question as to whether residual uncertainty after model fitting allowed for very different model predictions of intervention effect. Bayesian inference was made possible within Stan by introducing auxiliary prevalence variables and using an algebraic condition to fix these to the equilibrium solutions of the model. While Stan is capable of numerically solving ordinary differential equations, our algebraic log-likelihood allowed far more efficient (and in our experience) more reliable inference for a parameter space of this number of dimensions. Our approach may be applicable more widely in cases where a fit to an assumed equilibrium or steady decline through sparse data is sought.

Fitting our model under an assumption of equilibrium is a weakness of our study. Inevitably, TB epidemiology is not at equilibrium in any setting. However, our choice is defensible given that typical TB dynamics exhibit only single-digit annual percentage declines, and given a single prevalence survey and the substantial technical advantages discussed above bought by this assumption. Our model did not consider other subgroups that tend to mix non-randomly in real societies. Notably, we did not consider age; simultaneous patterns of non-random mixing by age and sex, together with non-uniform TB burden by age may nuance our findings. We also did not explicitly model HIV, whose prevalence is often unequal by sex. Such differences only appear in our model as averaged effects on progression in each sex. We identified little evidence to quantify our relative progression by sex, and adopted a relatively large uncertainty in this parameter (*α*) to allow fits to settings with large M:F ratios. The large uncertainty in the prior for *α* partially explains the dominance of this parameter in the M:F uncertainty analysis (Figure 3). Finally, we only considered a simple hypothetical intervention around equalizing case detection. Modelled impact may still depend on residual uncertainty for other types of intervention. Future work should consider inclusion of these features we have simplified, and realistic policy options that target disparities in detection, or potentially modifiable risk factors among men.

In conclusion, our analysis showed that assortative mixing does play a role in sex disparities in TB burden, and that this is particularly important under invasion dynamics. We found that equalizing case detection between the sexes alone is insufficient to eliminate disparities in burden. However, in settings with suboptimal case detection and large sex disparities in TB prevalence, this potentially achievable goal could still achieve meaningful reductions in overall TB burden.

## Data Availability

Link to the model codes is presented in the manuscript

## Acknowledgements

DSA is supported under the EDCTP2 programme supported by the European Union (grant number RIA2016S-1632-TREATS). PJD is supported by a Career Development Award from the UK MRC (MR/P022081/1). This UK funded award is part of the EDCTP2 programme supported by the European Union.

## Author contributions statement

PJD conceived the experiments, DSA conducted the experiments and analysed the results. All authors critically reviewed and edited the manuscript.

## Additional information

The authors declare no competing interests.

