## Supplementary Material for "Assortative social mixing and sex disparities in tuberculosis burden"

#### Contents

**1 Background** **2**

1.1 Sex disparity of TB burden . . . . . 2

**2 Parameterisation** **2**

**3 Condition for equilibria** **4**

**4 Impact of Intervention** **4**

**5 Posterior estimates** **7**

**6 Posterior distribution of parameters and model outputs** **8**

### 1 Background

Overall, men bear higher burden of tuberculosis compared to women. Sex disparity in TB burden considerably varies between settings with the highest male :female ratio reported in Uganda and Viet Nam, while Ethiopia has the reported lowest. In this work, we provide an analysis of the role of assortativity, differential progression and detection on TB M:F ratio at the two extreme settings, Ethiopia (M:F=1.2) and Uganda M:F=4.5) (Figure 1).

#### 1.1 Sex disparity of TB burden

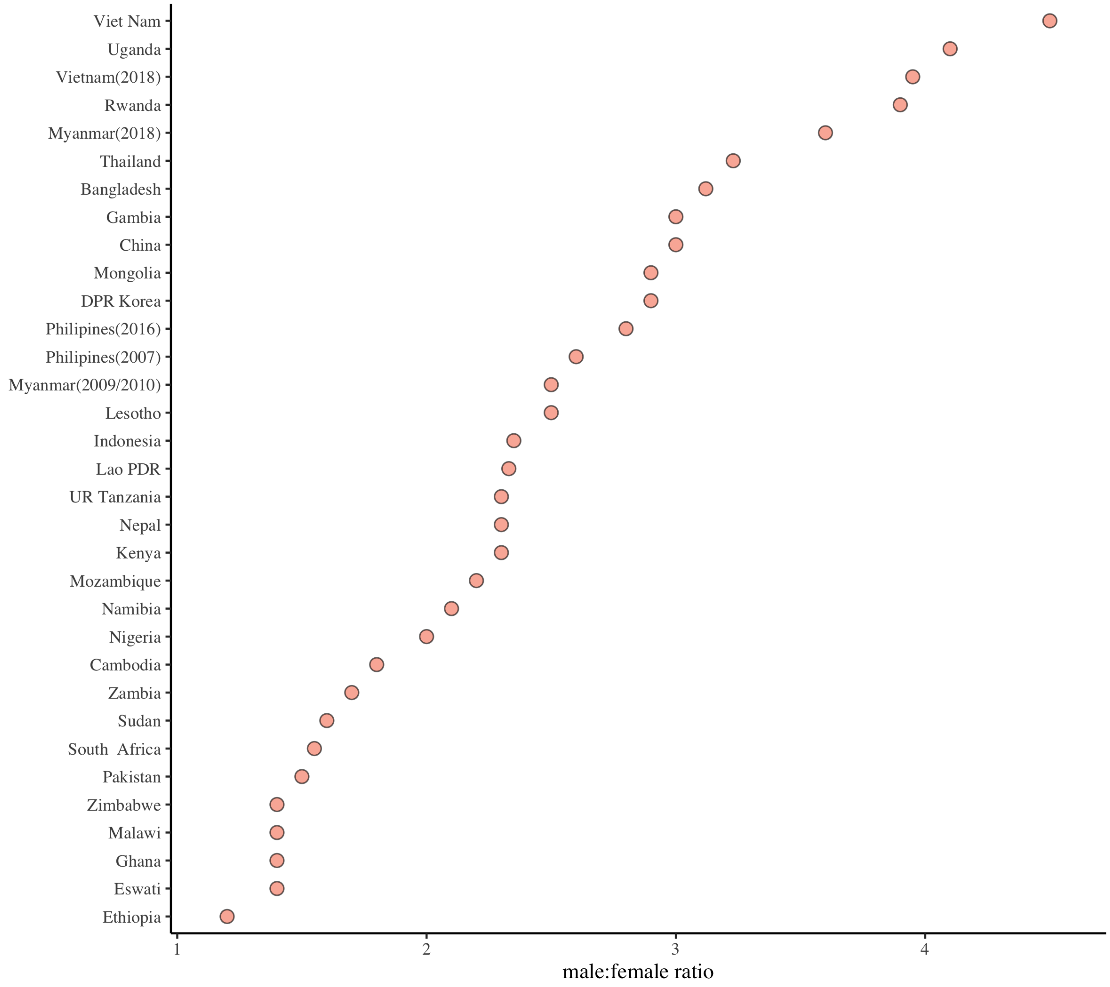

Figure 1: Sex disparity in TB prevalence (Adapted from WHO report, 2019)[1]

### 2 Parameterisation

This section provides brief descriptions of selected parameters in the model. In this study, we explored three mechanisms that could contribute to sex disparities in TB prevalence. In particular, we modelled the impact of social mixing, differential disease progression and detection.

**Social mixing**

To understand the contribution of social mixing on TB M:F ratio, we used a single scaling parameter that takes spectrum of values from -1(mixing only with opposite sex) to 1 (mixing only with the same sex). This parameter is expressed in terms of the proportion of contacts that are with the same sex (m) as :

$$\rho = 2m - 1, \tag{1}$$
$$m \sim \ LN(-0.57, 0.085)[2] \tag{2}$$

**Case detection rate**

The model is simulated using case detection proportion parameterised as an informative prior parameterised as Beta$(5.6, 3.5)$ based on WHO report for all TB types[**?**]. case detection proportion sampled from this prior is converted into case detection rate to be used in a model:

$$\delta = \frac{p}{1-p}(\gamma + \mu + \mu_t) \tag{3}$$

Case detection rates ($\delta$) generated this way are average rates and sex-stratified case detection rates are generated as follows:

$$\delta_f = \frac{2\delta}{1+\pi}$$
$$\delta_m = \pi \times \delta_f$$

where $\pi$- differential case detection, $\delta_f$- case detection rate in females and $\delta_m$-case detection rates in men.

**TB progression parameters**

TB progression parameters such as fast progression rate ($\epsilon$), reactivation rate ($\nu$) and relapse ($\omega$) are not available in sex stratification in literature. Therefore, values from literature on these parameters are considered as average and sex stratified rates were generated using similar approaches used for case-detection rates above.

For example, sex stratified reactivation rates were generated in our model as follows

$$\upsilon_f = \frac{2\upsilon}{1+\alpha}$$
$$\upsilon_m = \alpha \times \upsilon_f$$

where $\upsilon_f$ and $\upsilon_m$ are reactivation rates in men and women respectively, $\upsilon$ is the average reactivation rate, and $\alpha$ is the hazard ratio of progression in males compared to females.

**TB mortality and self-recovery rates**
As we are not modelling by smear status in this study, we generated sex specific average mortality rates by taking the average of TB mortalities in smear positive and smear negative cases. Similarly, sex specific TB self-recovery rates were also generated by taking the average of smear positive and smear negative recovery rates in within each sex group. A log-normal prior was set around these parameters to use in our model[3].

**Global and sex specific parameters**

In this model, only five parameters are sex specific: infectious proportion, fast progression, reactivation, relapse and case detection rate. In addition,potential differential transmission risk is captured by the social mixing parameter. All other model parameters are identical for men and women.

**Uncertainties in model parameters**
All model model parameters were uncertain and specified via priors,within a Bayesian framework. However, fixed values were used for two parameters - natural mortality ($\mu$) and treatment success ($\theta$).

### 3 Condition for equilibria

As described in the main text, we solved for the equilibrium condition of the model ordinary differential equations, which yielded a quadratic equation for the equilibrium prevalence:

$$Q(d;\beta,\Psi) = Ad^2 + Bd + C = 0, \tag{4}$$

where $d = D/N$, $\beta$ is the effective contact rate and $\Psi$ represents the parameters other than $\beta$. The coefficients in Equation 4 are

$$A = (\beta^2)\mu(-1+\psi)((\epsilon+\mu+\mu_t+\gamma)(\mu+\omega)(\mu+\tau)+\delta((\mu^2)+\mu(\omega+\tau)-\omega\tau(-1+\theta)+\epsilon(\mu+\omega+\tau\theta)));$$

$$B = -(\beta\mu(\beta\epsilon(-1+\psi)(\mu+\omega)(\mu+\tau)-(\mu+\omega)((\mu^2)(-2+\psi)-\mu_t\nu+\epsilon(\mu(-2+\psi)+\mu_t(-1+\psi)-\nu-\gamma)+ \mu(\mu_t(-2+\psi)-\nu-2\gamma+\psi\gamma-\kappa-\mu_t\kappa-\nu*\kappa-\gamma\kappa)(\mu+\tau)+\delta(-((\mu^3)(-2+\psi))+\nu\omega\kappa+\nu\omega\tau+\omega\kappa\tau+(\mu^2)(\nu- (-2+\psi)\omega+\kappa+2\tau-\psi\tau)+\mu(\kappa\tau+\nu(\omega+\kappa+\tau)+\omega(\kappa+(-2+\psi)\tau(-1+\theta)))- \nu\omega\tau\theta+\nu\kappa\tau\theta-\omega\kappa\tau\theta+\epsilon(-((\mu^2)(-2+\psi))+(-1+\psi)\omega\tau(-1+\theta)+\nu(\omega+\tau\theta)+\mu(\nu+2\omega-\psi\omega+\tau-\psi\tau+\tau\theta)))));$$

$$C = \beta\mu(\mu+\omega)(\epsilon(\mu+\nu)+\nu\kappa)(\mu+\tau)-\mu(\epsilon+\mu+\kappa)(((\mu^2)+\mu_t\nu+ \mu(\mu_t+\nu+\gamma))(\mu+\omega)(\mu+\tau)+\delta(\mu+\nu)((\mu^2)+\mu(\omega+\tau)-\omega\tau(-1+\theta)));$$

### 4 Impact of Intervention

The intervention considered in this study is narrowing the sex disparity in TB case detection rates.

To asses the impact of narrowing case detection gaps on TB prevalence and M:F ratio in TB prevalence, we simulated the intervention at different regions of joint posterior distributions of selected parameter combinations.

The joint distributions (parameter combinations) considered include:

- Relative detection( $\pi$) and relative progression ($\alpha$)
- Assortativity ( $\rho$) and relative progression ($\alpha$)
- Assortativity ( $\rho$) and relative detection ($\pi$)

#### 4.1 Sampling from different regions of posterior

We achieved sampling from three regions of joint posterior distributions by using ellipses with different radii and angle centred at different regions of the joint posterior. Data contained within each ellipse was extracted and used as input in a model used to simulate the impact of intervention. Data from different regions of parameter combinations were used to determine if the impact of an intervention is dependent on where the parameter is located in the parameter space (Figure 2).

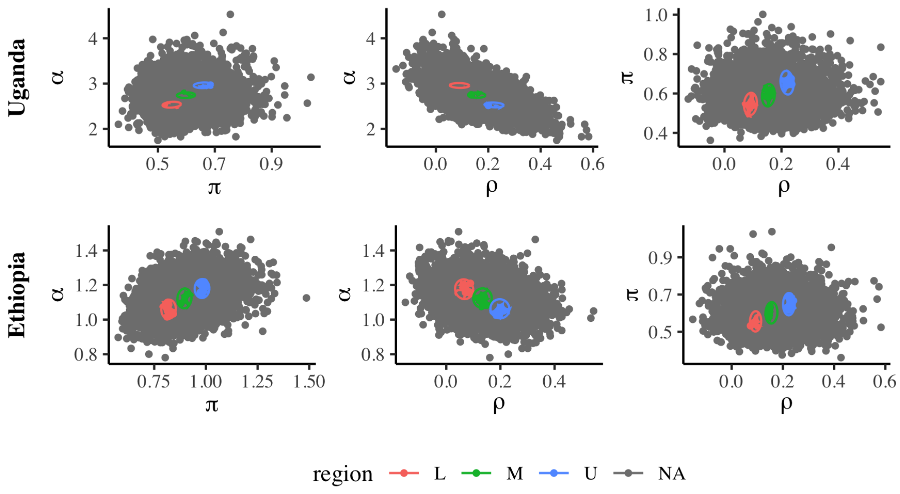

Figure 2: Sampled regions for the three selected parameter combinations

#### 4.2 Compatibility of sampled regions with data likelihood

Before the impact of intervention was simulated using parameters from the sampled regions, we assessed if parameters in the sampled region were supported by the data likelihood. This is achieved by comparing the log-posterior at these locations with the log-posterior from the whole parameter spaces of all parameters included in the model.

The plots below show that log-posteriors from the sampled regions coincide with the log-posterior from the entire parameter space. Columns **a, b** and **c** represent different parameter combinations: Parameters along column **a** represent combinations of $\pi$ (differential detection) and $\alpha$ (differential progression). Similarly, while those along column **b** represent $\rho$ (assortativity) and and $\alpha$, those under column **c** are for joint posterior distributions of $\rho$ and $\pi$ (Figure 3).

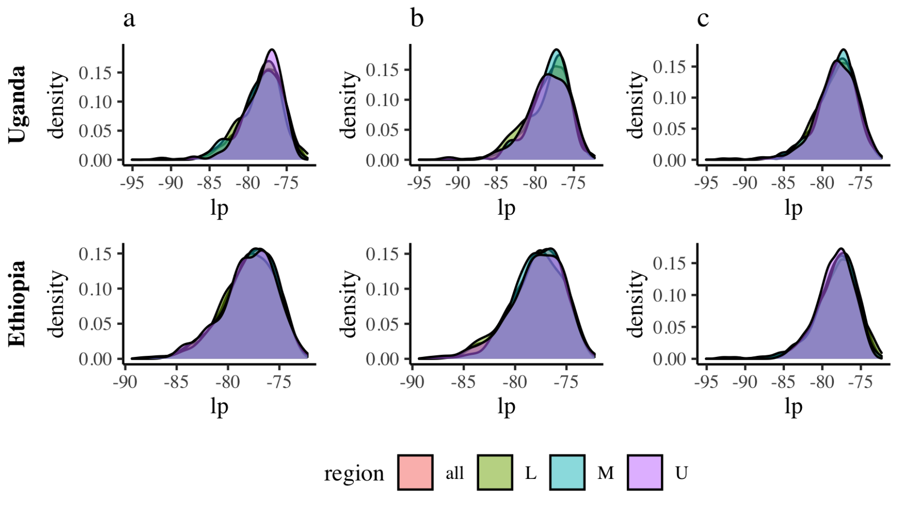

Figure 3: Log-posterior density at sampled regions

.

The impact of intervention was simulated at sampled regions of the selected joint distribution.

##### 4.3 Disease dynamics under intervention

The below plot shows the dynamics of TB under intervention using 200 parameters sets sampled from the whole region (Figure 4). In a high M:F ratio setting, the intervention can reduce prevalence to some extent. In the low M:F ratio setting,however, the prevalence remains unaffected with intervention. Similarly, eliminating case detection gap can meaningfully narrow the M:F ratios in a high M:F ratio setting , although its impact on low M:F setting is limited.

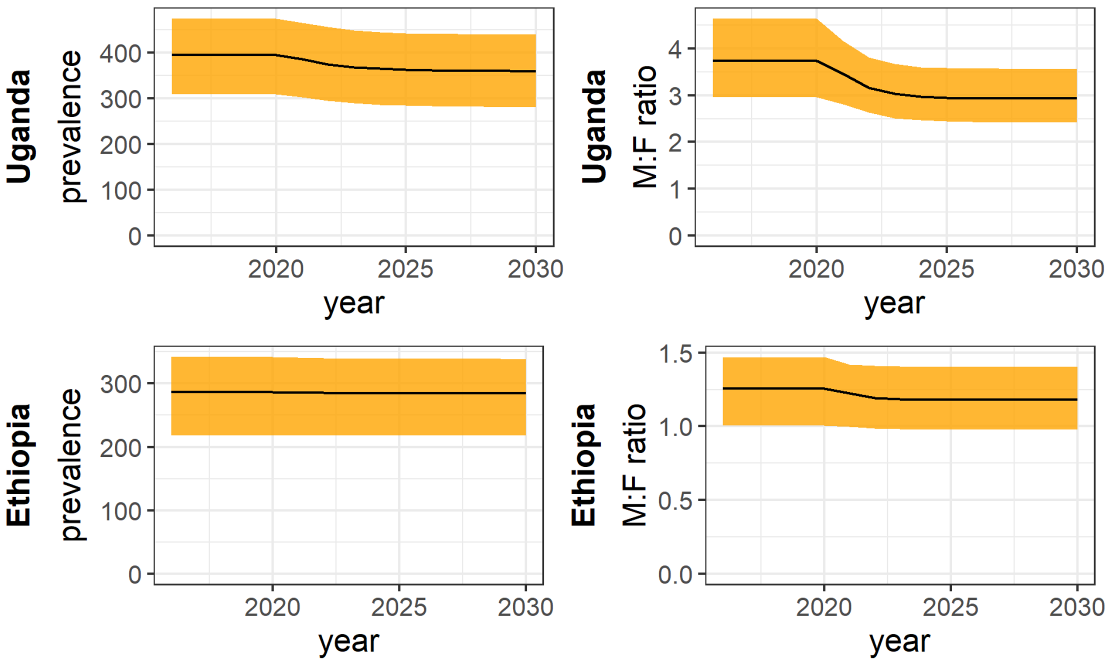

Figure 4: Disease dynamics under intervention

##### 4.4 Pairs plots

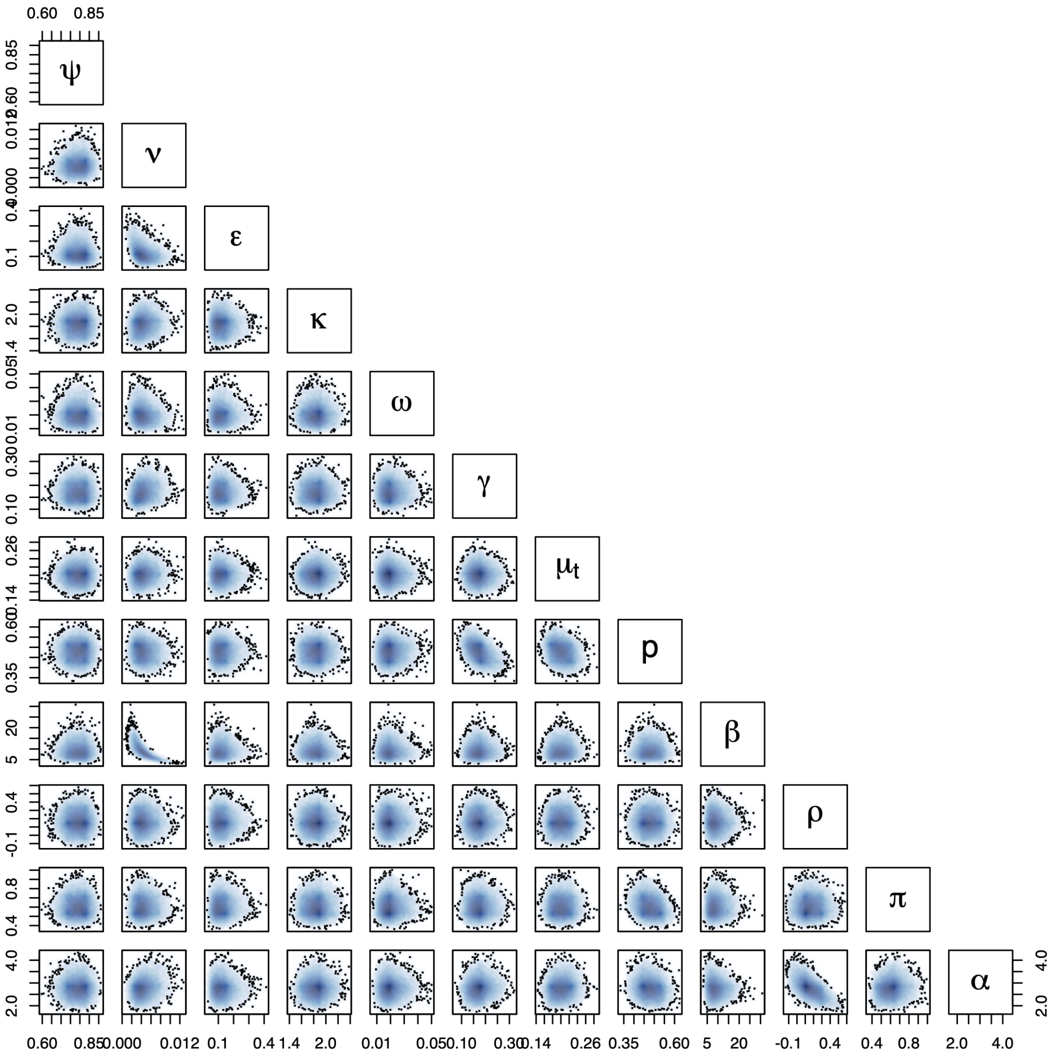

Figure 5: correlation beteween model parameters-Uganda

### 5 Posterior estimates

The following table compares posterior estimates from the two exemplar settings. Our posterior estimates from the two exemplar settings were identical for most of the parameters involved. The estimates differed only with respect to the proportion of cases detected, differential detection and differential disease risk (Table 1).

NB: The $\pi$ (differential detection) compares the hazard of males being detected compared to women. Similarly, $\alpha$ (differential risk of progression) measures disease progression in men as compared with women.

Table 1: Posterior parameter estimates in the two exemplar settings

| Parameters | Description | Ethiopia: 95% CrI | Uganda: 95 % CrI |
|---|---|---|---|
| $\psi$ | Partial protection | 0.79(0.70, 0.86) | 0.79(0.70, 0.86) |
| $\gamma$ | Natural recovery, year$^{-1}$ | 0.18(0.12, 0.25) | 0.17(0.12, 0.24) |
| $\epsilon$ | Fast progression rate, year$^{-1}$ | 0.12(0.05, 0.22 ) | 0.12(0.06, 0.22 ) |
| $\kappa$ | Stabilisation rate, year$^{-1}$ | 1.8(1.6, 2.1) | 1.8(1.6, 2.1) |
| $\nu$ | Reactivation rate, year$^{-1}$ | 0.005(0.002, 0.009) | 0.004(0.002, 0.008) |
| $\omega$ | Relapse rate, year$^{-1}$ | 0.02(0.01, 0.03) | 0.02(0.01, 0.03) |
| $\mu_t$ | TB mortality rate, year$^{-1}$ | 0.20(0.17, 0.24) | 0.20(0.17, 0.24) |
| $\beta$ | Effective contact rate, year$^{-1}$ | 10(5.6, 17) | 9(4.9, 16) |
| $\rho$ | Assortative mixing | 0.14(-0.04, 0.34) | 0.16(-0.02, 0.36) |
| $p$ | Proportion of cases detected, year$^{-1}$ | 0.62(0.55, 0.68) | 0.48(0.40, 0.55) |
| $\pi$ | Relative detection rate, year$^{-1}$ | 0.90(0.69, 1.16) | 0.60(0.46, 0.78) |
| $\alpha$ | Relative progression rate,year$^{-1}$ | 1.1(0.95, 1.3) | 2.7(2.2, 3.4) |

### 6 Posterior distribution of parameters and model outputs

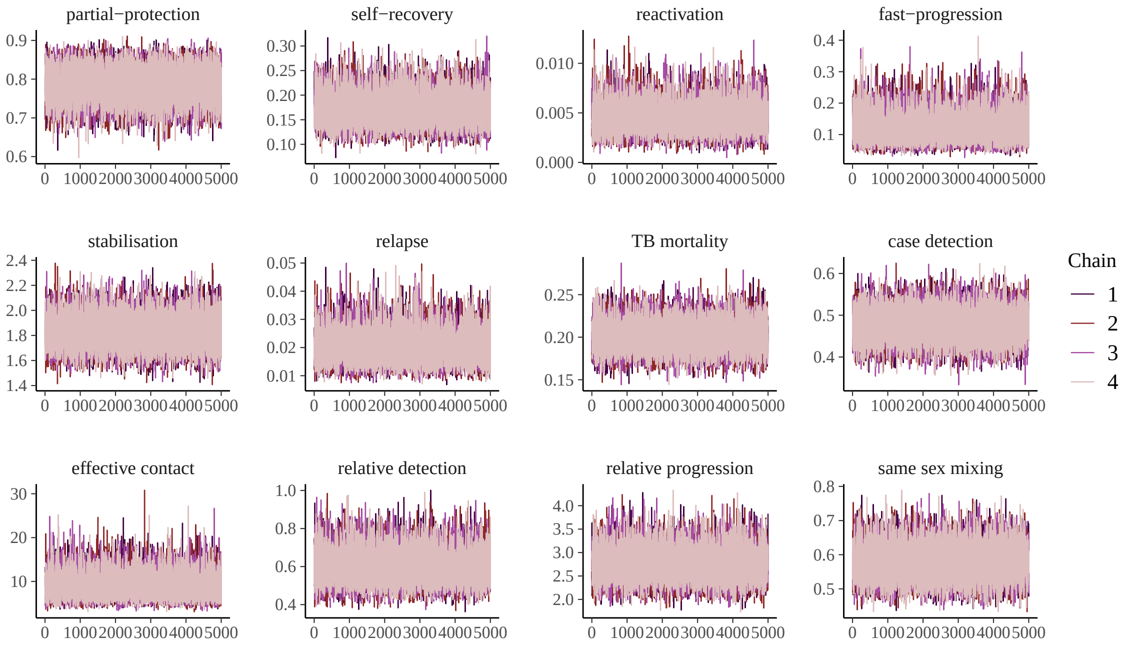

Figure 6: Trace plots of posterior estimates of parameters-Uganda

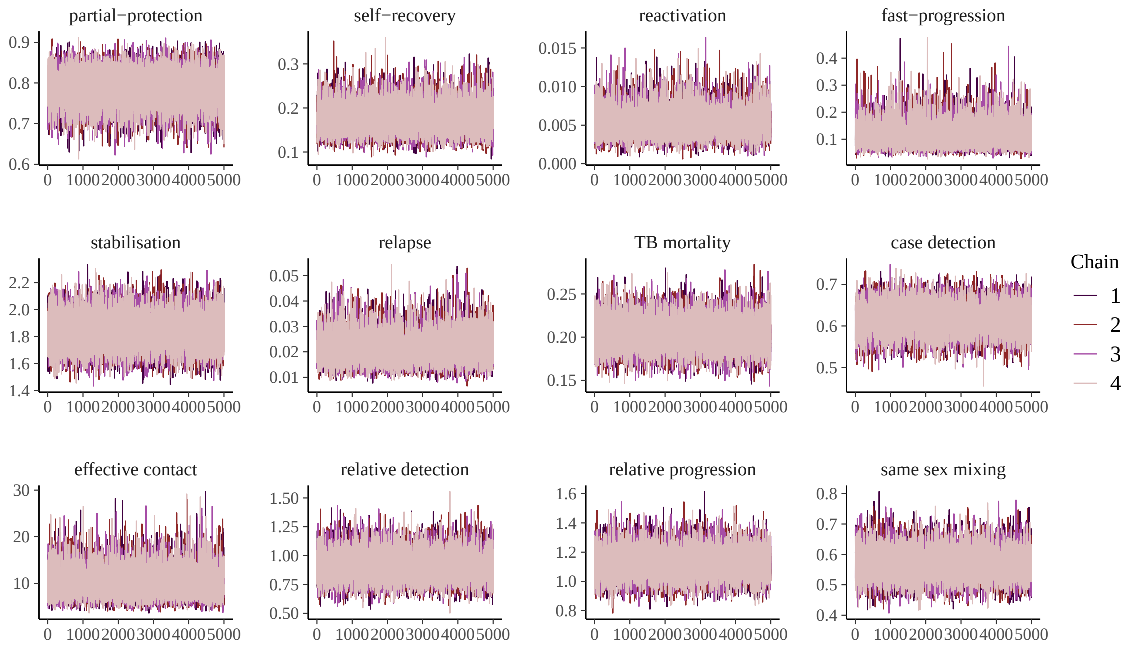

Figure 7: Trace plots of posterior estimates of parameters-Ethiopia

[3] Romain Ragonnet, Jennifer A Flegg, Samuel L Brilleman, Edine Tiemersma, Yayehirad A Melsew, Emma S McBryde, and James M Trauer. Revisiting the natural history of pulmonary tuberculosis: a bayesian estimation of natural recovery and mortality rates. *BioRxiv*, page 729426, 2019.
